# A PHILOSOPHICAL PERSPECTIVE ON NURSING INTERVENTION IN ELDERLY WITH DEPRESSION

**DOI:** 10.1101/2022.08.08.22278530

**Authors:** Enik Suhariyanti

## Abstract

**Introduction:** Depression is one of the most common mental illnesses in the elderly, can be found in various health care settings and is ranked as the fourth disease in the world as a cause of disability. This literature aims to study the philosophy of nursing intervention in elderly with depression. It viewed from three philosophical perspectives, namely ontology, epistemology, and axiology.

**Method:** This study uses a literature study design from 6 databases, namely: Science Direct, Pubmed, Proquest, Wiley, Sagepub, and Neliti research. The search used various keyword combinations with the help of Boolean operators, including: “Intervention” OR “Nursing Intervention AND “Elderly” AND “Depression”, This research is a quantitative study focusing on publications between 2017-2021. Thus, 18 eligible articles were obtained. Article quality is monitored using the CASP checklist. The results of the articles obtained come from several countries including Indonesia.

**Result:** Nursing interventions include three categories, first, physical/exercise therapy, psychological therapy, and spiritual therapy to reduce depression levels in the elderly in various settings such as nursing homes, correctional facilities, and other general communities including home visits, using individual and group approaches.

**Conclusion:** The results of this review provide an explanation that several interventions that can be carried out in the elderly can save costs, are feasible and easy to do, and are effective for reducing depression levels in the elderly, and can improve quality of life, overcome other psychological problems.

## INTRODUCTION

The elderly are one of the vulnerable groups (Kiik, Sahar, & Permatasari, 2018). With increasing age, physiological functions decrease due to the aging process so that many non-communicable diseases appear in the elderly and degenerative problems reduce the body’s resistance so that the elderly are susceptible to infectious disease infections (Infodatin, 2016). Elderly also often experience social problems, in the form of isolation from society due to decreased physical function experienced, for example reduced hearing sensitivity, as well as ways of speaking that are sometimes incomprehensible (Azizah, 2011). The elderly are very vulnerable to personality changes, role changes and changes in interests related to retirement age, loss of a partner (Stanley, Mickey, & Patricia Gauntlett Beare, 2007).

Psychosocial changes in the elderly due to depression are very detrimental to the health of the elderly, both for physical and mental health (Putra, 2014). The problem of depression in the elderly requires serious treatment because it can have a broad impact on health and life. Depression in the elderly will affect the physical activity and quality of life of the elderly (de Oliveira, Souza, Rodrigues, Fett, & Piva, 2019). Elderly who experience depression can cause changes in physical, thought, feeling, and behavior so that the elderly tend to have a low quality of life (Utami, Liza, & Ashal, 2018).

Philosophy is an attitude or view of life and an applied field to help individuals to evaluate their existence in a more satisfying way. Philosophy leads to understanding and understanding leads to appropriate action. In general, philosophical systematics has three main discussions or sections, namely; epistemology or theory of knowledge which discusses how to acquire knowledge, ontology or theory of nature which discusses the nature of everything that gives birth to knowledge and axiology or theory of value which discusses the use of knowledge. Thus, studying these three branches is very important in understanding philosophy which is so broad in scope and discussion.

## METHOD

The research method used is an integrative literature review. This review proposes a better understanding of the effectiveness of nursing interventions against depression in the elderly. We systematically searched Science Direct, Pubmed, Proquest, Wiley, Sagepub, and Neliti research. The search used various keyword combinations with the help of Boolean operators, including: “Intervention” OR “Nursing Intervention AND “Elderly” AND “Depression. The inclusion criteria applied in this study were peer-reviewed articles in English that discussed nursing interventions against depression in the elderly. Articles published in the last five years (2017-2021). The quality of the study was carried out using the CASP checklist (Critical Appraisal Skills Program, 2018). Researchs studies are conducted in various fields that specifically examine nursing interventions against depression in the elderly.

This study is a quantitative study with a randomized controlled trial (RCT) and full text method. We used the PRISMA Flowchart 2009 (Moher et al., 2010) to record the article review and inclusion process (see Figure 1). The first search with keywords found 39,008 articles. related. Then the search was limited to the 2017-2021 range and got 37,358 articles, restrictions were made to keep the author current based on the latest research results. After reading the articles obtained, the researchers divided them into 2 criteria, namely inclusion and exclusion so that the articles were eligible. The inclusion criteria, namely the researcher took the original article using the systematic review method, RCT, and quasy experiment got 6,035 articles, which discussed specifically related to depression in the elderly and not accompanied by other comorbidities, got 101 articles. While the exclusion criteria are articles that duplication and non-pharmacological interventions. From all literature searches, 39,008 articles were obtained, then 38,907 articles were excluded and the remaining 101 were included. Then the researchers searched for 81 duplicate articles, and there were 2 non-pharmacological articles so that there were 18 articles that matched were selected for final inclusion (Figure 1).

**Figure 1.**
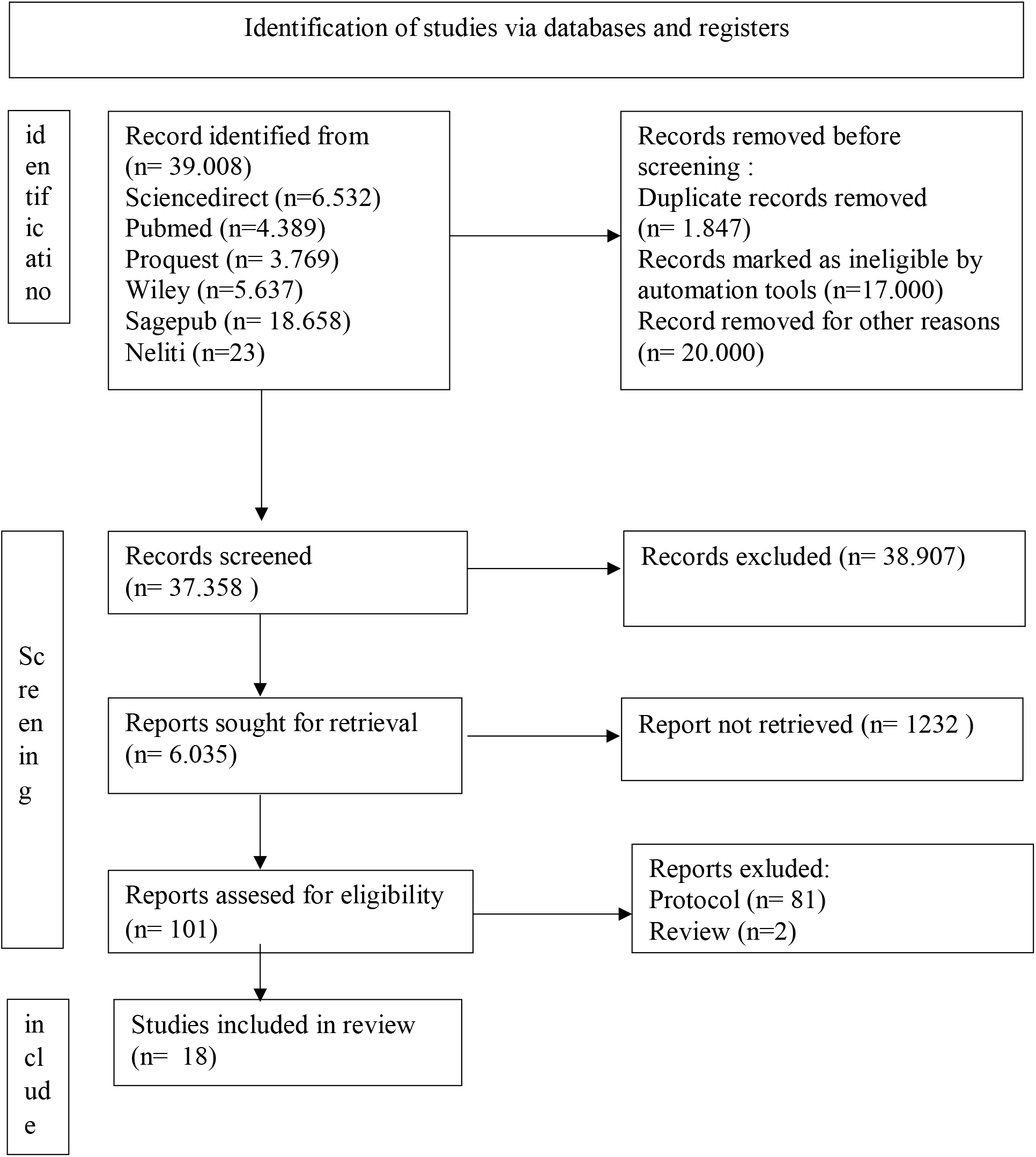
PRISMA Flowchart of Literature Search and Screening Process

## RESULT AND DISCUSSION

### 1. Ontology Study on the nursing intervention in elderly with depression

Depression is one of the most common mental illnesses in the elderly, ranking the fourth disease in the world as a cause of disability. The situation in which a person feels a loss of enthusiasm for activities that are often carried out beyond the time limit of 14 consecutive days can be said to be an early symptom of depression (Vahia, 2013). It is estimated that by 2020, depression will increase and may be ranked second in world health problems (WHO, 2017). Depression in the elderly is caused by several factors including the loss of a partner, chronic physical illness, the use of several drugs, the presence of psychological stressors. (Varma, 2012) Depression can affect an individual’s ability to function optimally and risk of suicide (WHO, 2017). This is also supported by several research results which say that 33.3% of the elderly are depressed with 23.3% of mild depression and 10% of the elderly experiencing severe depression (Mutiara, Rinita, & Purnama, 2019). But other studies say that most of the elderly depression rates are within normal limits of 46.9% because the elderly receive good support from their families (Livana, Susanti, Darwati, & Anggraeni, 2018).

Psychosocial changes in the elderly due to depression are very detrimental to the health of the elderly, both for physical and mental health (Putra, 2014). The problem of depression in the elderly requires serious treatment because it can have a broad impact on their health and life. Depression in the elderly will affect the physical activity and quality of life of the elderly (de Oliveira, Souza, Rodrigues, Fett, & Piva, 2019). Elderly who experience depression can cause changes in physical, thinking, feeling, and behavior so that the elderly tend to have a low quality of life (Utami, Liza, & Ashal, 2018). Therefore, its are interested in conducting research to overcome the problem of depression in the elderly.

Nursing intervention is an action that includes made to assist the individual (client) in moving from the current level of health to the desired level of expected outcomes. Types of Nursing Interventions in dealing with the elderly with depression are independent nursing interventions is an action that nurses take on clients independently without an active role from other health workers and collaborative nursing interventions are interventions carried out by nurses on patients or clients in the form of collaboration with other health workers.

Interventions carried out to treat depression in the elderly can be grouped into three categories. First, psychological therapy such as Reminiscence (Hong Kong, Indonesia, India, Iran), Art Therapy (Brazil), Therapy using pets (Italy), Psychoeducation (Spain). Second, physical therapy/exercise such as Qigong and Tai Chi (Hong Kong), Yoga (India), (Australia), Light Therapy (China), Multidomain Intervention/physical activity, healthy diet, social activity, and brief cognitive restructuring (Republic of Korea). Third, spiritual therapy such as listening to the Koran recital (Indonesia), Spiritual-Religious Psychotherapy (Iran).

### 2. Epistemological Study on the nursing intervention in elderly with depression

This intervention includes non-pharmacological methods that can be applied to treat depression in the elderly in various settings. Based on the analysis of the article above, it is very effective for the elderly to be at least 60 years old who are depressed so that the quality of life of the elderly remains optimal. The quality of life of the elderly as an assessment that is very dependent on individual subjectivity about his position in life in the context of the cultural and value system in which the elderly lives and is related to the goals, expectations, standards and concerns they experience (World Health Organization, 2014; Xavier, Ferraz, Marc, Escosteguy, & Moriguchi, 2003).

Previous research stated that the elderly who live with their families have independence, are still quite able to meet all their own social and economic needs and are directly involved in activities with their families. This proves that the role of the family will make the elderly experience positive changes in their lives (Putri, Fitriana, Ningrum, & Sulastri, 2015). This condition is also supported by the results of research that there is a relationship of social interaction with the quality of life of the elderly in relation to the adjustment of the elderly which can affect current or future life (Andesty & Syahrul, 2018; Indrayani & Ronoatmodjo, 2018).

The importance of the active role of the family in doing therapy for the elderly. Where family support is very important for the elderly because lack of family support can trigger depression, such as feelings of not getting attention, loneliness, feeling useless, and will only be a burden to the family. Changes experienced by the elderly both physically, psychologically, socially, and spiritually will affect the occurrence of depression problems so that in terms of providing intervention/therapy to overcome depression problems can be grouped into three categories, namely physical, psychological, and spiritual therapy so that the life and health of the elderly can be improved. optimal.

### 3. Axiologi Study on the nursing intervention in elderly with depression

From the literature review activities of journals it was shows the synthesis of the study population based on the articles reviewed. The articles obtained came from several countries, from 18 eligible articles, the research was conducted in Indonesia, Australia, Spain, India, Hong Kong, Brazil, Italy, China, Republic of Korea, and Iran. This intervention can be carried out in various settings such as nursing homes, correctional facilities, and other general communities including home visits. This intervention can also be carried out using an individual or group approach. Eligibility criteria stated for respondents in This study is the elderly who do not have mental health problems. The sample size in this study varied, it could be done on elderly men and women, the average age was at least 60 years.

Interventions carried out to treat depression in the elderly can be grouped into three categories. First, psychological therapy such as Reminiscence (Hong Kong, Indonesia, India, Iran), Art Therapy (Brazil), Therapy using pets (Italy), Psychoeducation (Spain). Second, physical therapy/exercise such as Qigong and Tai Chi (Hong Kong), Yoga (India), Baduanjin Qigong (China), Arm Therapy (Australia), Light Therapy (China), Multidomain Intervention/physical activity, healthy diet, social activity, and brief cognitive restructuring (Republic of Korea). Third, spiritual therapy such as listening to the Koran recital (Indonesia), Spiritual-Religious Psychotherapy (Iran). The instruments used to measure the level of depression in the elderly used GDS (Geriatric Depression Scale), MADRS (Montgomery-Asberg Depression Rating Scale), DASS (Depression Anxiety and Stress Scales), Goldberg’s General Mental Health Questionnaire, and BDI (Beck Depression Inventory).

## CONCLUSION

This review can answer the research objective, which is to find some literature that can be given to reduce the level of depression in the elderly in various settings such as nursing homes, correctional facilities, and the elderly who live with their families. Overall, the intervention is simple, feasible, and cost-effective, as well as non-pharmacological to reduce the level of depression in the elderly. This intervention can also be carried out individuals and groups. Other interventions can also improve quality of life, reduce anxiety levels, and other psychological problems faced by the elderly.

## Data Availability

All data produced in the present work are contained in the manuscript

## ACKNOWLEDGEMENTS

I would like to thank all those who have supported the completion of this article

## REFERENCES

Ambrosi, C., Zaiontz, C., Peragine, G., Sarchi, S., & Bona, F. (2018). Randomized controlled study on the effectiveness of animal-assisted therapy on depression, anxiety, and illness perception in institutionalized Elderly. Psychogeriatrics, 19(1), 55–64. https://doi.org/10.1111/psyg.12367

Andesty, D., & Syahrul, F. (2018). Hubungan interaksi sosial dengan kualitas hidup lansia di Unit Pelayanan Terpadu (UPTD) Griya Werdha Kota Surabaya Tahun 2017. The Indonesian Journal of Public Health, 13(2), 169–180. https://doi.org/10.20473/ijph.vl13il.2018.169-180

Azizah. (2011). Keperawatan Lanjut Usia. Yogyakarta: Graha Ilmu.

Casañas, R., Martín Royo, J., Fernandez-San-Martín, M. I., Raya Tena, A., Mendioroz, J., Sauch Valmaña, G., … Martín Lopez, L. M. (2019). Effectiveness of a psychoeducation group intervention conducted by primary healthcare nurses in patients with depression and physical comorbidity: Study protocol for a randomized, controlled trial. BMC Health Services Research, 19. https://doi.org/10.1186/s12913-019-4198-7

Centre for Reviews and Dissemination. (2008). Systematic Reviews: CRD’s guidance for undertaking reviews in health care. York, UK: York University: Centre for Reviews and Dissemination University of York.

Chobe, S., Chobe, M., Metri, K., Patra, S. K., & r, N. (2020). Impact of yoga on cognition and mental health among elderly: A systematic review. Complementary Therapies in Medicine, 52. https://doi.org/10.1016/j.ctim.2020.102421

Ciasca, E. C., Ferreira, R. C., Santana, C. L. A., Forlenza, O. V., Dos Santos, G. D., Brum, P. S., & Nunes, P. V. (2018). Art therapy as an adjuvant treatment for depression in elderly women: A randomized controlled trial. Revista Brasileira de Psiquiatria, 40(3), 256–263. https://doi.org/10.1590/1516-4446-2017-2250

Critical Appraisal Skills Programme. (2018). CASP checklist for systematic review. Middle Way Oxford. Retrieved from www.casp-uk.net

de Oliveira, L. D. S. S. C. B., Souza, E. C., Rodrigues, R. A. S., Fett, C. A., & Piva, A. B. (2019). The effects of physical activity on anxiety, depression, and quality of life in elderly people living in the community. Trends in Psychiatry and Psychotherapy, 41(1), 36–42.https://doi.org/10.1590/2237-6089-2017-0129

Gill, B. K., Cant, R., Lam, L., Cooper, S., & Lou, V. W. Q. (2020). Non-pharmacological depression therapies for older Chinese adults: A systematic review & meta-analysis. Archives of Gerontology and Geriatrics, 88. https://doi.org/10.1016/j.archger.2020.104037

Higgins, J. (2008). Assessing risk of bias in included studies. In J. P. Higgins & D. G. Altman (Eds.), Cochrane Handbook for Systematic Reviews of Interventions (pp. 1–50). Chicester, UK: John Wiley &Sons Ltd.

Indrayani, & Ronoatmodjo, S. (2018). Faktorfaktor yang berhubungan dengan kualitas hidup lansia di Desa Cipasung Kabupaten Kuningan Tahun 2017. Jurnal Kesehatan Reproduksi, 9(1), 69–78. https://doi.org/10.22435/kespro.v9i1.892.69-78

Infodatin. (2016). Situasi Lanjut Usia di Indonesia. Kementerian Kesehatan Republik Indonesia.

Jing, L., Jin, Y., Zhang, X., Wang, F., Song, Y., & Xing, F. (2018). The effect of Baduanjin qigong combined with CBT on physical fitness and psychological health of elderly housebound. Medicine, 97(51), 1–7. https://doi.org/10.1097/MD.0000000000013654

Juniarni, L., Lindayani, L., Nurdina, G., & Hendra, A. (2020). Efektifitas terapi reminiscence terhadap tingkat depresi pada lansia. Jurnal Ilmu Keperawatan Jiwa, 3(4), 411–420.

Kemenkes. (2017). Situasi Lansia DiIndonesia Tahun 2017.

Kemenkes Republik Indonesia. (2020). HINDARI LANSIA DARI COVID 19. http://www.padk.kemkes.go.id/article/read/2020/04/23/21/hindari-lansia-dari-covid-19.html

Kiik, S. M., Sahar, J., & Permatasari, H. (2018). Peningkatan kualitas hidup lanjutusia (Lansia) di kota Depok dengan latihan keseimbangan. Jurnal Keperawatan Indonesia, 21(2), 109–116.https://doi.org/10.7454/jki.v21i2.584

Klompstra, L., Ekdahl, A. W., Krevers, B.,Milberg, A., & Eckerblad, J. (2019).Factors related to health-related quality of life in older people with multi morbidity and high health care consumption over a two-year period. BMC Geriatrics, 19(1), 1–8. https://doi.org/10.1186/s12877-019-1194-z

Livana, Susanti Y., Darwati, L. E., & Anggraeni, R. (2018). Gambaran Tingkat Depresi Lansia. Jurnal Keperawatan Dan Pemikiran IlMiah,80–93.

Mehra, A., Rani, S., Sahoo, S., Parveen, S., Singh, A. P., Chakrabarti, S., & Grover, S. (2020). A crisis for elderly with mental disorders: Relapse of symptoms due to heightened anxiety due to COVID-19. Asian Journal of Psychiatry, 51(April), 102114. https://doi.org/10.1016/j.ajp.2020.102114

Miller, K. J., Gonçalves-Bradley, D. C., Areerob, P., Hennessy, D., Mesagno, C., & Grace, F. (2020). Comparative effectiveness of three exercise types to treat clinical depression in older adults: A systematic review and network meta-analysis of randomised controlled trials. Ageing Research Reviews, 58. https://doi.org/10.1016/j.arr.2019.100999

Musavi, M., Mohammadian, S., & Mohammadinezhad, B. (2017). The effect of group integrative reminiscence therapy on mental health among older women living in Iranian nursing homes. Nursing Open, 4(4), 303–309.https://doi.org/10.1002/nop2.101

Mutiara, A., Rinita, A., & Purnama, D. N. (2019). Gambaran tingkat depresi pada lansia di wilayah kerja Puskesmas Guguak Kabupaten 50 Kota Payakumbuh. Health & Medical Journal, 1 (2), 12–16. https://doi.org/10.33854/heme.v1i2.235

Pramesona, B. A., & Taneepanichskul, S. (2018). The effect of religious intervention on depressive symptoms and quality of life among indonesian elderly in nursing homes: A quasi-experimental study. Clinical Interventions in Aging, 13, 473–483. https://doi.org/10.2147/CIA.S162946

Putra, H. (2014). Pengaruh Terapi Reminiscence (Mengenang Masa Lalu yang Menyenangkan) terhadap Depresi pada Lansia di Unit Rehabilitasi Sosial Pucang Gading Semarang 2014

Putri, S. T., Fitriana, L. A., Ningrum, A., & Sulastri, A. (2015). Studi komparatif: Kualitas hidup lansia yang tinggal bersama keluarga dan panti. Jurnal Pendidikan Keperawatan Indonesia, 1(1), 1. https://doi.org/10.17509/jpki.v1i1.1178

Ramanathan, M., Bhavanani, A., & Trakroo, M. (2017). Effect of a 12-week yoga therapy program on mental health statusin elderly women inmates of a hospice. International Journal of Yoga, 10(1), 24.https://doi.org/10.4103/0973-6131.186156

Roh, H. W., Hong, C. H., Lim, H. K., Chang, K. J., Kim, H., Kim, N. R., … Son, S. J.(2020). A 12-week multidomain intervention for late-life depression: A community-based randomized controlled trial. Journal of Affective Disorders, 263, 437–444. https://doi.org/10.1016/j.jad.2019.12.013

Rokayah, C., Kusnandar, K., & Putri, M. H. (2019). Pengaruh terapi reminiscence terhadap penurunan tingkat depresi pada lansia. Jurnal Imliah Permas: Jurnal Imliah STIKES Kendal, 9(2), 73–78.

Sedaghat Ghotbabadi, S., & Haji Alizadeh, K. (2018). The effectiveness of spiritual-religion psychotherapy on mental distress (depression, anxiety and stress) in the elderly living in Nursing Homes. Health, Spirituality and Medical Ethics, 5(1), 20–25. https://doi.org/10.29252/jhsme.5.1.20

Sollami, A., Gianferrari, E., Alfieri, E., Artioli, G., & Taffurelli, C. (2017). Pet therapy: An effective strategy to care for the elderly? An experimental study in a nursing home. Acta Bio-Medica : Atenei Parmensis, 88(1), 25–31.https://doi.org/10.23750/abm.v88i1

Stanley, Mickey, & Patricia Gauntlett Beare. (2007). Buku Ajar Keperawatan Gerontik (2nd ed.). Jakarta: EGC.

Tarugu, J., Pavithra, R., Vinothchandar, S., Basu, A., Chaudhuri, S., & John, K. R. (2019). Effectiveness of structured group reminiscence therapy in decreasing the feelings of loneliness, depressive symptoms and anxiety among inmates of a residential home for the elderly in Chittoor district. International Journal Of Community Medicine And Public Health, 6(2), 847–854. https://doi.org/10.18203/2394-6040.ijcmph20190218

Utami, A. W., Liza, R. G., & Ashal, T. (2018). Hubungan kemungkinan depresi dengan kualitas hidup pada lanjut usia di Kelurahan Surau Gadang Wilayah Kerja Puskesmas Nanggalo Padang. Jurnal Kesehatan Andalas, 7(3), 417. https://doi.org/10.25077/jka.v7i3.896

Vahia, V. (2013). Diagnostic and statistical manual of mental disorders 5: A quick glance. Indian Journal of Psychiatry, 55(3), 220. https://doi.org/10.4103/0019-5545.117131

Varma, G. S. (2012). Depression in the Elderly : Clinical Features and Risk Factors. Aging and Disease, 3(6), 465–471.

WHO. (2017). Depression and Other Common Mental Disorders: Global Health Estimates. https://doi.org/CC BY-NC-SA 3.0 IGO

World Health Organization. (2014). WHOQOL: Measuring quality of life. Retrieved August 3, 2020, from https://www.who.int/healthinfo/survey/whoqol-qualityoflife/en/

World Health Organization. (2018). Ageing and health.

Xavier, F. M. F., Ferraz, M. P. T., Marc, N.,Escosteguy, N. U., & Moriguchi, E. H.(2003). Elderly people’s definition of quality of life. Revista Brasileira de Psiquiatria, 25(1), 31–39.https://doi.org/10.1590/S1516-44462003000100007

Yuan, Y., Li, J., Jing, Z., Yu, C., Zhao, D.,Hao, W., & Zhou, C. (2020). The role of mental health and physical activity in the association between sleep quality and quality of life among rural elderly in China: A moderated mediation model. Journal of Affective Disorders, 273(May), 462–467. https://doi.org/10.1016/j.jad.2020.05.093

Zhao, X., Ma, J., Wu, S., Chi, I., & Bai, Z. (2018). Light therapy for older patients with non-seasonal depression: A systematic review and meta-analysis. Journal of Affective Disorders, 232, 291–299. https://doi.org/10.1016/j.jad.2018.02.041

